# Evolution and impact of COVID-19 outbreaks in care homes: population analysis in 189 care homes in one geographic region

**DOI:** 10.1101/2020.07.09.20149583

**Authors:** Jennifer K Burton, Gwen Bayne, Christine Evans, Frederike Garbe, Dermot Gorman, Naomi Honhold, Duncan McCormick, Richard Othieno, Janet Stevenson, Stefanie Swietlik, Kate E Templeton, Mette Tranter, Lorna Willocks, Bruce Guthrie

## Abstract

**Background:** COVID-19 has had large impact on care-home residents internationally. This study systematically examines care-home outbreaks of COVID-19 in a large Scottish health board.

**Methods:** Analysis of testing, cases and deaths using linked care-home, testing and mortality data for 189 care-homes with 5843 beds in a large Scottish Health Board up to 15/06/20.

**Findings:** 70 (37.0%) of care-homes experienced a COVID-19 outbreak, 66 of which were in care-homes for older people where care-home size was strongly associated with outbreaks (OR per 20-bed increase 3.50, 95%CI 2.06 to 5.94). There were 852 confirmed cases and 419 COVID-related deaths, 401 (95.7%) of which occurred in care-homes with an outbreak, 16 (3.8%) in hospital, and two in the 119 care-homes without a known outbreak. For non-COVID related deaths, there were 73 excess deaths in care-homes with an outbreak, but no excess deaths in care-homes without an outbreak, and 24 fewer deaths than expected of care-home residents in hospital. A quarter of COVID-19 related cases and deaths occurred in five (2.6%) care-homes, and half in 13 (6.9%) care-homes.

**Interpretation:** The large impact on excess deaths appears to be primarily a direct effect of COVID-19, with cases and deaths are concentrated in a minority of care homes. A key implication is that there is a large pool of susceptible residents if community COVID-19 incidence increases again. Shielding residents from potential sources of infection and rapid action into minimise outbreak size where infection is introduced will be critical in any wave 2.

**Funding:** Not externally funded.

**Research in context**

**Evidence before this study**
We searched PubMed and the medRxiv preprint server using terms ‘long-term care’, ‘nursing home’, ‘care home’, or ‘residential care’ combined with ‘COVID-19’ and/or ‘SARS-CoV-2’, updated to 25^th^ June. The existing published literature highlights the large impact in care-homes, and that atypical disease presentation, asymptomatic carriage and a presymptomatic infectious period is common in both residents and staff. One living systematic review confirms the international outbreak burden among residents and staff and high but varied international mortality rates. International modelling studies have failed to take account of the care-home environment and context, making estimates informed by general community transmission of infection. Only one peer-reviewed study was identified which evaluated US nursing home characteristics associated with outbreaks, finding associations with larger facility size, urban location, and ethnicity, but no association with quality ratings or ownership.

**Added value of this study**
This study reports data for all 189 care homes in one large Scottish health board, where 37% experienced an outbreak of COVID-19, with 95% of outbreaks in care-homes for older people. The number of beds was the only care-home characteristic statistically significantly associated with the presence of an outbreak. One-third of affected care homes had only single cases or short outbreaks, but sustained outbreaks were common, and there was evidence of potential reintroduction of infection in some care-homes with >14 day gaps between confirmed cases. Cases and mortality were heavily concentrated. In care-homes with an outbreak there were 472 excess deaths (12.3% of bed capacity, 3.1 times the average in the previous five years), 85% of which were COVID-19 related. There were only 16 COVID-19 related deaths and 14 other deaths of care-home residents in hospital in the same period, consistent with ∼20 non-COVID excess deaths occurring in care-homes being deaths that would have happened anyway. 99% of the excess deaths and of the COVID-19 related deaths were in care-homes with an outbreak, suggesting that quality and safety of care in the wider care system was not affected.

**Implications of all the available evidence**
Outbreak patterns varied considerably and more detailed understanding of why some care homes avoided or controlled outbreaks would allow learning to prepare for wave two. Systematic, regular testing and use of whole genome sequencing will inform understanding of transmission dynamics and future outbreak management. Future research should consider the built environment and organisation of care as other potentially modifiable factors to support infection control. Improving national and local data on the care-home population is a priority both for COVID-19 and for ensuring this vulnerable population receives better care in the future.

## Background

Internationally, institutional care settings for older adults have experienced high rates of COVID-19 infection and mortality among residents and staff.^1^ In the UK, such settings are usually called care-homes, providing 24-hour nursing and/or residential care for those whose complex needs cannot be accommodated at home or in other settings.^2^ In the UK and other countries including the US, care-home funding is often a mix of self-pay and state funding, and the care sector has been under increasing financial and capacity strain because of population ageing and constraints on public funding.^3^ Robust national data on the care-home population are lacking and data sources are fragmented, meaning our understanding of the needs and outcomes of residents is poor.^4,5^

Early epidemiological data identified very high mortality from COVID-19 in some care-home settings, for example affecting 33% of residents in a single US facility.^6^. Atypical presentation of COVID-19 is prevalent, with delirium, postural instability and diarrhoea common presenting symptoms without fever or cough.^7,8^ The role of pre-symptomatic and asymptomatic transmission has become clearer over time, with one US study finding that 56% of positive residents asymptomatic when tested and the majority of asymptomatic residents then developing symptoms in the subsequent four days.^9^ In four London care-homes with large outbreaks where all residents and staff were tested twice one week apart, 40% of residents tested positive, of whom 43% were asymptomatic at the time of testing, and 18% had atypical symptoms. Four percent of staff were positive, all of whom were asymptomatic, and viral genome sequencing found evidence of multiple introductions of infection.^10^

Deaths in care-homes from COVID-19 account for 47% of all COVID-19 mortality in Scotland,^11^ but residents dying in hospital have been separately reported to date. National data are aggregated which means our understanding of variation between care-homes is limited. The aim of this study is to describe the evolution of outbreaks of COVID-19 in all care-homes in one large health board in Scotland, including timing of outbreaks, number of confirmed cases in residents, care-home characteristics associated with the presence of an outbreak, and deaths of residents in both care-homes and hospitals.

## Methods

### Setting

NHS Scotland has taxpayer funded universal coverage healthcare with no patient fees or copayments for medical care. NHS Scotland health boards have responsibility for delivery of all NHS care in their geographical area (including public health). State social care funding is means-tested and either funded by local authorities, or self-funded with some state funding for personal or nursing care. Social care provision for older people is primarily delivered by private providers. The health board examined is home to ∼1 million people,^12^ of whom∼16% are aged 65 and over and ∼7% aged 75 and over.^13^ There are four Integration Joint Boards (IJBs) which commission care-home services and are coterminous with the local authority areas in the region. Key milestones in national policy and the local Public Health response to COVID-19 are summarised in box 1.

### Data sources

Local Public Health data on COVID-19 resident testing results were linked to publicly available data on Care Home Services, collated by the national regulator (the Care Inspectorate) and published in April 2020.^14^ Analysis is of data for 188 registered care-home services and one NHS run short-stay and respite facility, and the linked Public Health data is complete to 15/06/20.

Deaths of residents in care-homes were identified from National Records Scotland (NRS) death registration defined as deaths with an institutional code compatible with a care home, with manual verification and allocation to a specific care-home to ensure accuracy. Deaths of care-home residents in hospital were identified from NRS death registration where the place of death was a hospital, with addresses linked to Care Inspectorate care-home registered addresses using postcode and text matching. Death data is complete to 07/06/20.

### COVID-19 outbreak definition

Following current practice in the public health team, the start of an outbreak was defined as the date that the first resident in a care-home has a positive COVID-19 test using regional virology laboratory polymerase chain reaction (PCR) testing of nasopharyngeal swabs.

### Other variables

Care Inspectorate data used included (1) Care-home type (recategorised as older people, other adult services [combining physical/sensory impairment, alcohol and drugs, mental health, respite care, blood borne viruses], learning disabilities, children and young people; (2) Number of beds with number of registered places used as a proxy if not available; (3) Risk Assessment Document (RAD) score (CI) which is Care Inspectorate assessment determining intensity of independent inspection (low, medium and high risk); (4) Ownership (private, local authority, and voluntary or not-for-profit); (5) Locality (the IJB/local authority the care-home is located in).

Public health outbreak reports were used to count how many outbreaks of any infectious disease (most commonly norovirus, influenza and scabies) had been reported to public health since March 2014 (dichotomised into ≥5 outbreaks in the past six years) as a measure of historical infection control practice.

Death was examined in terms of week of death registration which is more stable over recent short periods of time than date of death. Registration is legally required to be within eight days of death, although most deaths are registered within three days. COVID-related deaths were defined as any death where there was a record of confirmed (ICD-10 code U07.1) or suspected (U07.2) COVID-19 in any position on the death certificate. All other deaths were defined as non-COVID related deaths.

### Statistical methods

Logistic regression was used to estimate univariate and adjusted odds ratios of the presence of an outbreak by care-home characteristics, with primary analysis restricted to older people’s care homes and sensitivity in all care homes. The evolution of the epidemic at care-home and resident level was examined descriptively, and the Estimated Dissemination Ratio (EDR) in care-homes calculated as the number of cases in a seven day period divided by the number of cases in the preceding seven days. EDR requires no assumptions about transmission routes or infectious periods, and gives a direct assessment of the slope of the epidemic curve and how that is changing.^15^ EDR has similar interpretation to R0, with an EDR>1 indicating acceleration and EDR<1 indicating slowing. Excess deaths (COVID-related and non-COVID related) were estimated by comparing observed deaths to the average in the same period over the last five years, in all care-homes, and separately in those with and without an outbreak. The first positive COVID-19 test in a care-home resident in week 12 (beginning 16/03/20), and excess deaths were therefore estimated from week 13 (beginning 23/03/20) onwards.

### Findings

There were 189 care-homes included, 109 (57.7%) services for older people, 14 (7.4%) for other adults, 26 (13.8%) for learning disabilities, and 40 (21.2%) for children and young people. These 189 care-homes had 5843 beds, 5227 (89.5%) of which were in care-homes for older people.

### Evolution of the epidemic at resident and care-home level

The first resident test was carried out on 10^th^ March 2020, and the first positive test was in week 12 (beginning 16/03/20) (Figure 1, panel A). There were a total of 55 outbreaks in the five subsequent weeks to 19/04/20. Over the next six weeks to 31/05/20, there a further 15 more sporadic outbreaks, with no outbreaks in the final two weeks of observation.

**Figure 1:**
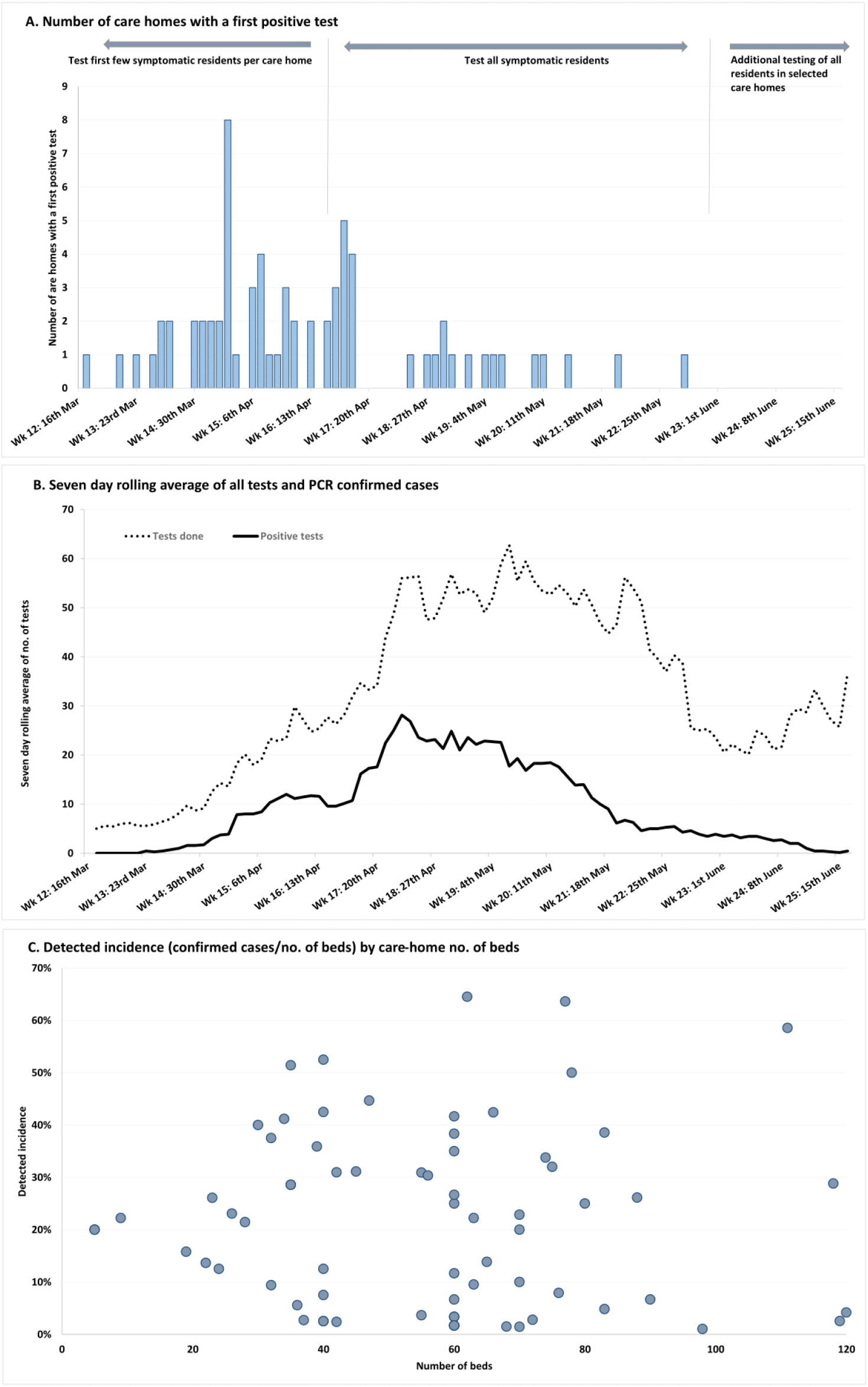
Evolution of outbreaks over time

The number of residents tested per day rose rapidly, peaking at 50-60 in late April (when testing policy changed from ‘first few symptomatic cases in each care-home’ to ‘all symptomatic cases’) to late May, then fell, with a later rise when testing policy changed to ‘test all residents in care-homes with ongoing outbreaks’. The seven day moving average of numbers of residents with a confirmed positive case rose rapidly, peaking at 28 per day on 23^rd^ April 2020 (Figure 1, panel C). Daily averages have fallen continually since then. Mirroring this, the Estimated Dissemination Rate was 7.9 on 03/04/20 falling to 0.8 on 16/04/20. There was a second peak of 2.8 on 23/04/20 and a steady decline to below one from 28/04/20 with values below one persistently since then (Supplementary Figure 1).

Fifty-one (72.9%, 95%CI 60.7 to 82.5) care-homes with an outbreak had one or more residents with negative tests before their first confirmed case (Table 1, Figure 2). The number of positive cases per care-home ranged from 1 to 65 (median 7, IQR 2 to 17). Twelve (17.1%) care-homes only had a single case, and 14 (20.3%) had 2-4 cases. A quarter (26.6%) of all cases were in just five care-homes, and almost half (47.8%) were in 12 care-homes.

**Table 1:** Outbreaks in 70 care-homes

|  | No. (% , 95% CI) of care-homes |
| --- | --- |
| Negative testing before first positive test |  |
| One or more negative tests in care-home | 51 (72.9%, 60.7 to 82.5) |
| First test done in care-home was positive | 19 (27.1%, 17.5 to 39.3) |
| No. of positive cases |  |
| 1 | 12 (17.1%, 9.5 to 28.4) |
| 2-4 | 14 (20.0%, 11.7 to 31.6) |
| 5-9 | 11 (15.7%, 8.5 to 26.8) |
| 10-19 | 17 (24.3%, 15.2 to 26.3) |
| 20-29 | 10 (14.3%, 7.4 to 25.2) |
| 30+ | 6 (8.6%, 3.5 to 18.4) |
| Detected incidence (cases per bed) |  |
| 0 to 9.9% | 24 (34.3%, 23.6 to 46.7) |
| 10 to 19.9% | 8 (11.4%, 5.4 to 21.8) |
| 20 to 29.9% | 16 (22.9%, 14.0 to 34.7) |
| 30 to 39.9% | 10 (14.3%, 7.4 to 25.2) |
| 40 to 49.9% | 6 (8.6%, 3.5 to 18.4) |
| 50% and over | 6 (8.6%, 3.5 to 18.4)) |
| Ongoing outbreaks (evaluated 16//06/20) |  |
| No positive cases for $\geq 28$ days | 43 (61.4%, 49.0 to 72.6) |
| No positive cases for 14-27 days | 18 (25.7%, 16.3 to 37.8) |

**Figure 2:**
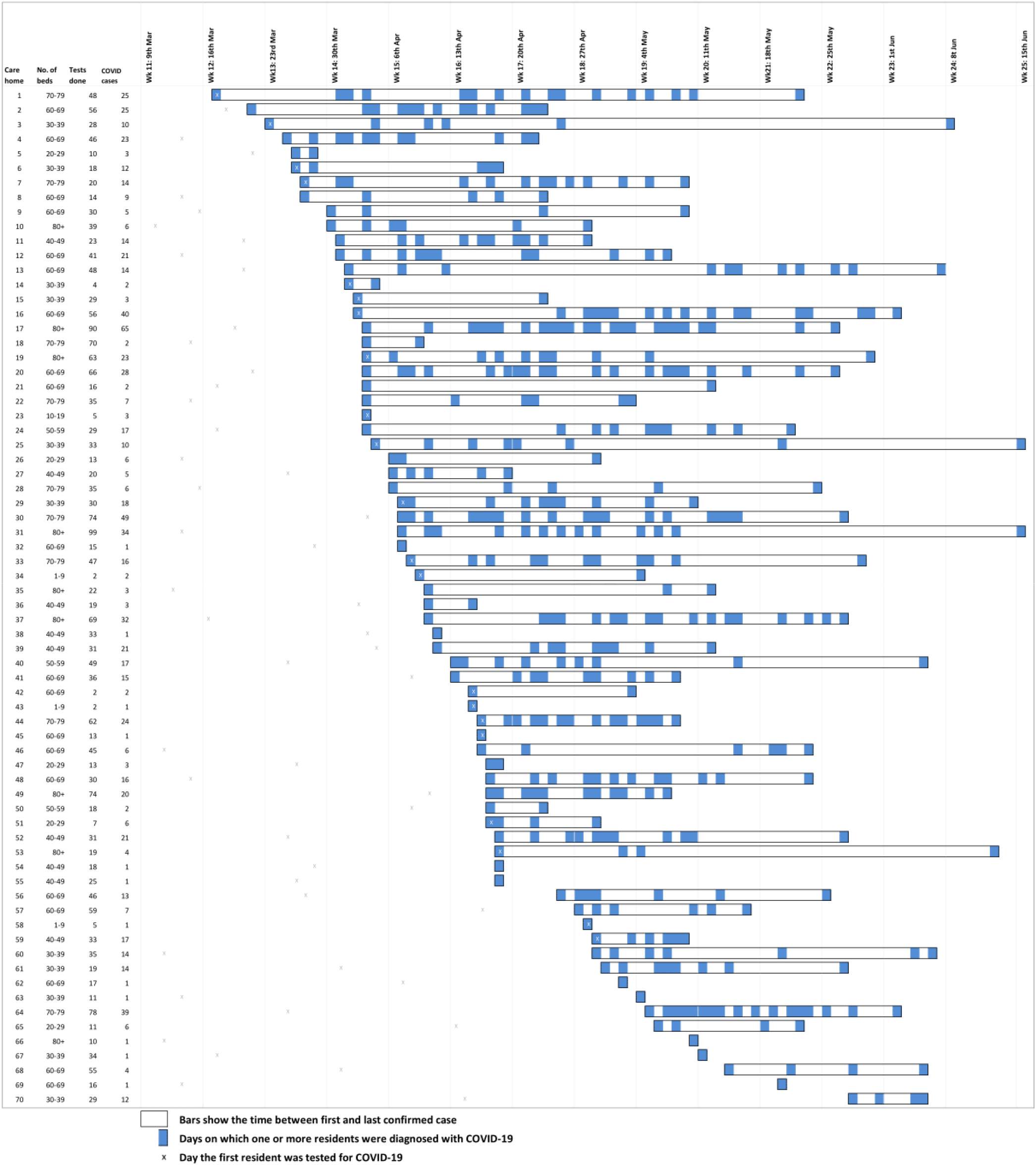
Patterns of outbreak for each care-home with an outbreak

The detected incidence (positive cases per bed) in care-homes ranged from 1.0% to 66.7% (median 20.7%, IQR 6.4% to 34.1%) but there was no relationship between care-home number of beds and detected incidence (Figure 1, panel C; Pearson-R 0.02, p=0.90). There was considerable heterogeneity in patterns of diagnosed infection, with 13 (18.6%) care-homes only having cases diagnosed on a single day, and a further 10 (14.3%) only having cases over 2-14 days (Figure 2). Although there was evidence of sustained infection over several weeks in many care homes, in 18 (25.7%) there were gaps between diagnosed cases of 14-27 days, and in 5 (7.1%) gaps of ≥28 days, consistent with new introductions of infection (Figure 2, care-homes 3, 21, 31, 35, 53). However, 43 (61.4%) care-homes had not had a new case for at least 28 days consistent with outbreaks ceasing (and a further 18 (25.7%) had not had a new case for 14-27 days).

### Care home characteristics associated with an outbreak

Seventy (37.0%, 95%CI 30.2 to 44.4) care-homes had an outbreak (one or more positive tests). Sixty-six (94.3%) outbreaks were in older people’s care-homes, where 60.6% had experienced an outbreak compared to six (5.0%) of all other care-home types (Table 2). Care home characteristics systematically varied by care-home type. Older people’s care-homes were much larger (median 48 beds vs eight for all other types combined), more likely to be in private ownership (67.9% vs 30.0%) or to have a RAD score of medium or high risk (41.3%% vs 15.2%) or to have a history of multiple prior outbreaks of other infectious disease (28.4% vs 0%), and less likely to be located in locality 4 (14.7% vs 36.3%).

**Table 2:** Care homes with an outbreak by care-home type

|  | Median (range)<br>no. of beds | No resident testing<br>done<br>No. (%) of care-homes | One or more<br>residents tested, but<br>no positive tests<br>No. (%) of care-homes | One or more<br>residents with a<br>positive test<br>(outbreak)<br>No. (%) of care-homes |
| --- | --- | --- | --- | --- |
| Older people (n=109) | 42 (10-119) | 4 (3.7) | 39 (35.8) | 66 (60.6) |
| Other adult (n=14)* | 8 (4-60) | 5 (37.5) | 8 (57.1) | 1 (7.1) |
| Learning disabilities (n=26) | 5 (4-11) | 13 (50.0) | 11 (42.3) | 2 (7.7) |
| Children & young people (n=40) | 5 (1-45) | 32 (80.0) | 7 (17.5) | 1 (2.5) |
| All care-homes (n=189) | 23 (1-119) | 54 (28.6) | 65 (34.4) | 70 (37.0) |

\* Physical/sensory impairment, alcohol and drugs, mental health, short-break and respite care (including an NHS-run service), blood borne viruses

**Table 3:** Older people’s care-home characteristics associated with an outbreak

| Care home characteristics (no. of care-homes, no. of beds) | No. (%) with an outbreak<br>N=109 care homes | Univariate OR (95% CI) | Adjusted OR (95% CI)* |
| --- | --- | --- | --- |
| No. of beds (per 20 bed increase) | - | 3.50 (2.06 to 5.94) | 3.50 (2.06 to 5.94) |
| Regulatory risk assessment score<sup>#</sup> |  |  |  |
| Low (n=64, 2767 beds) | 38 (59.4) | Reference | - |
| Medium (n=24, 1153 beds) | 12 (50.0) | 0.68 (0.27 to 1.76) |  |
| High (n=21, 1307 beds) | 16 (76.2) | 2.19 (0.71 to 6.72) |  |
| Ownership |  |  |  |
| Private (n=74, 3835 beds) | 43 (58.1) | Reference | - |
| Local authority (n=19, 795 beds) | 13 (68.4) | 1.56 (0.54 to 4.56) |  |
| Voluntary or not for profit (n=16, 597 beds) | 10 (62.5) | 1.20 (0.40 to 3.66) |  |
| 5+ other types of outbreak in last 6 years |  |  |  |
| No (n=78, 3197 beds) | 41 (52.6) | Reference | - |
| Yes (n=31, 2030 beds) | 25 (80.6) | 3.76 (1.39 to 10.2) |  |
| Locality |  |  |  |
| Locality 1 (n=65, 3169 beds) | 42 (64.6) | Reference | - |
| Locality 2 (n=17, 654 beds) | 8 (41.7) | 0.49 (0.17 to 1.43) |  |
| Locality 3 (n=11, 554 beds) | 7 (63.6) | 0.96 (0.25 to 3.62) |  |
| Locality 4 (n=16, 850 beds) | 9 (56.3) | 0.70 (0.23 to 2.14) |  |

<sup>#</sup> Care Inspectorate (Scottish Care Home Regulator) risk assessment score determined on independent inspection
\* No other care home characteristic is statistically significant once number of beds is accounted for

In univariate analysis for older people’s care homes only, there were statistically significant associations between the presence of an outbreak and number of beds (OR per 20-bed increase 3.50), a history of multiple previous outbreaks (OR 3.76), and regulatory risk assessment score (OR high-risk vs low 2.19). However, in adjusted analysis, only number of beds (OR per 20-bed increase 3.50, 95%CI 2.06 to 5.94 per 20-bed increase). Similar results were found in sensitivity analysis of all care-homes.

### Deaths of care-home residents in care-homes and in hospital

There were 419 deaths of care-home residents where COVID-19 was recorded on the death certificate, 403 (96.2%) occurring in care-homes and 16 (3.8%) occurring in hospital. The number of COVID-19 deaths rose rapidly to peak at 63-68 per week in weeks 16 to 19 (weeks beginning 13/04 to 04/05).

Almost all COVID-19 related deaths occurring in care-homes were in those with an outbreak (401 deaths, representing 99.5% of COVID-19 related deaths occurring in care-homes). In the 70 care-homes with an outbreak, there were 472 excess deaths in weeks 13-20 (67.6% of all deaths in these care-homes in that time), of which 399 (84.5%) were COVID-19 related. In care-homes with an outbreak, the peak ratio of observed to expected deaths was 5.3 in week 17 (figure 3, panel A).

**Figure 3:**
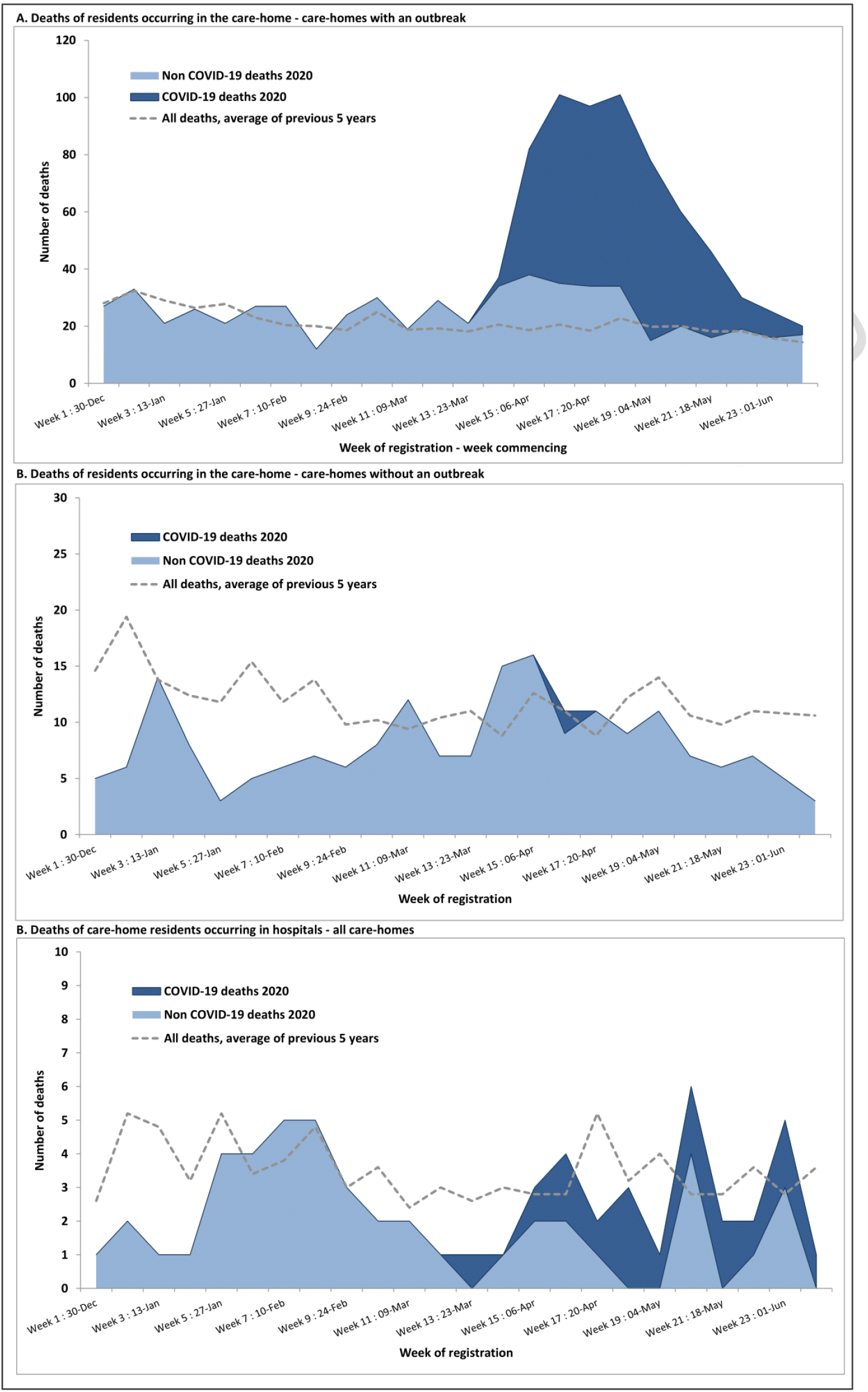
deaths in care-homes Source: National Records Scotland (NRS) death registration. Data is by week of registration (which is later than date of death). NRS data is provisional and may be subject to change, particularly later data if registration is delayed

In comparison, in the 119 care-homes without an outbreak, there was no clear excess mortality and only two COVID-19 related deaths (one stated as confirmed and one as suspected on the death certificate, both untested and not known to public health) (figure 3, panel B).

In 2014-2019, 10.2% of deaths of care-home residents were in hospital. Sixteen care-home residents died of COVID-19 in hospital in weeks 13-20 (3.8% of all care-home resident COVID deaths), with 14 non-COVID deaths, compared to an average in the same period of 39.2 over the previous five years.

## Discussion

Just over one-third of care-homes in the health board had experienced a COVID-19 outbreak, but with wide variation in the size, duration and pattern of outbreaks. Almost all outbreaks were in care-homes for older people but the only care-home characteristics statistically significantly associated with the presence of an outbreak was number of beds. Many care-homes experienced single cases or short outbreaks, but sustained or repeated outbreaks were common. In the period examined, there were 852 confirmed cases and 399 COVID-19 related deaths recorded in 70 care-homes with 3451 beds, plus an estimated additional 73 (15.5%) excess deaths not recorded as COVID-19 related. A quarter of confirmed cases and deaths were in just five care-homes, and approximately half in twelve care-homes. In the remaining 119 care-homes, there were no outbreaks, no excess deaths, and only two COVID-19 related deaths reported neither of whom were tested.

There were only 16 (3.8%) COVID-19 related deaths of care-home residents in hospital, and approximately one-third the expected non-COVID-19 deaths in hospitals during the epidemic, consistent with some of the observed non-COVID-19 excess deaths in care homes being deaths that would otherwise have occurred in hospital (although numbers are small).

Strengths include the inclusion of all care-homes in a geographical area using linked data, and examination of resident deaths in care-homes and in hospitals. Availability of resident testing was higher and earlier than in many other areas because the regional virology laboratory had capacity, and a public health outreach team supported rapid testing as part of an active outbreak response programme. Limitations include that testing policy and therefore case ascertainment changed over time. Before 17/03/20, testing typically ceased after two to five cases were positive (except where outbreaks were large when additional testing was sometimes done). Subsequently all suspected cases were tested, with asymptomatic resident testing implemented from the end of May in care-homes with ongoing outbreaks. Case numbers are likely to be underestimated, particularly early in the epidemic.^10^ However, the absence of excess mortality in care-homes without an outbreak makes it unlikely that any large outbreaks were missed. Finally, the study examines 189 care-homes and so is relatively underpowered to examine associations with care-home characteristics.

Data from the Care Quality Commission shows that 36% of English care-homes report an outbreak compared to 37% in this study.^16^ A larger proportion of all COVID-19 deaths are reported to have occurred in Scottish compared to English care-homes (47% vs 28%),^11,17^ although both figures are in the range reported internationally (from 24% in Hungary to 82% in Canada).^1^ This may reflect differences in admission practices in England and Scotland, since approximately one-eighth of COVID-19 related deaths of care home residents in England are in hospital^18^ versus only 3.8% in this study. Data from a London point-prevalence study of four nursing homes for ∼400 residents identified a 26% mortality rate across the homes, higher than that observed here, likely reflecting selection of care-homes with large outbreaks in London.^10^ US data has also shown significant associations between the presence of outbreaks and larger care-home size, and lack of association with regulatory quality ratings.^19^

The impact of COVID-19 on care-homes has been very large, but this study highlights that the direct and indirect impact on mortality is concentrated in homes with an outbreak. In this study, almost all excess deaths were in the one-third of care-homes with outbreaks, with further concentration in this group since approximately half of confirmed cases and deaths were in just 7% of care-homes. Equally, the observed pattern of outbreaks highlights that any increase in community transmission has the potential to drive further outbreaks, because the majority of older people’s care homes have not had an outbreak, or rapidly controlled their outbreak, meaning that large numbers of residents likely remain susceptible. Predictive modelling of risk must be specific to the care-home population, rather than based on community data.^20^

Approximately one in six excess deaths were not attributed to COVID-19 on the death certificate, but these deaths were almost exclusively in care-homes with an outbreak, and there was no evidence of significant numbers of COVID-19 or other deaths in hospital. This suggests that excess deaths in care-home residents in this health board are not related to wider changes to the system of care (e.g. inappropriate withdrawal of primary care or inappropriate reductions in hospital admission affecting all care-home residents). Whether excess deaths not attributed to COVID-19 in care-homes with an outbreak are a direct effect (undiagnosed COVID-19 in residents presenting with atypical symptoms) or an indirect effect (e.g. related to changes in care for uninfected residents when staff are overwhelmed and/or short-staffed) needs investigation because at least some may be preventable.

The analysis highlights that univariate associations with care-home characteristics will often be misleading because care-home type (the population served), ownership, regulatory quality ratings and a history of multiple previous outbreaks of other infectious diseases are all strongly associated with care-home size, which (unsurprisingly) dominates associations with the presence of a COVID-19 outbreak.^19^ Although care-home size cannot be altered without losing places for existing residents, there may be potential to create discrete units within care-homes where smaller numbers of staff and residents are effectively cohorted to create self-contained units. Such efforts will be complicated by individual care-home built environment, and will be difficult to sustain without rapid outside support during any large COVID-19 outbreak when staff illness and absence may risk compromising safe care. Additional measures to respond to new outbreaks of COVID-19 will also be required, including maintaining high provision of adequate Personal Protective Equipment (PPE), better support for infection control, ensuring self-isolation and active surveillance of residents and staff to ensure early detection of outbreaks and ongoing transmission, and staffing support for care-homes with many staff absent (box 2). However, COVID-19 has high infectivity, which is reflected in high rates of nosocomial infection of other patients and staff in hospital settings despite better established PPE access and infection control training than social care settings.^21^ Infection control is therefore intrinsically difficult in care-home settings where a priority is to maintain social and cognitive function through interaction. Shielding residents and care-homes from further outbreaks will be essential but poses difficult problems, for example in relation to the impact of preventing or minimising family and friends visiting which has its own impact on quality of life.^22^

There are a number of unanswered questions. First, SARS-CoV-2 transmission dynamics in this context are not well understood, including the extent to which outbreaks are sustained by ongoing transmission within care-homes and/or by introduction of new infections, and the relative importance of staff introducing infection vs newly admitted residents (or visitors if care-homes open up to visiting). More systematic testing of both residents and staff with whole genome sequencing to trace transmission chains will be of great value, and likely inform public health response (both are underway in this health board).

Second, the care-home characteristics examined in this study are high-level. Research is needed to understand the relative importance of the built environment, the intensity and nature of staff-resident and resident-resident interactions (which will vary considerably depending on the client group), variations in use of agency staff and investment in staff training, access to and effective use of Personal Protective Equipment, and infection control procedures. Some of this data can be collected by survey, but some may require direct observation of care which is challenging during an epidemic. Third, research is needed to understand excess deaths, including exploring variation in the attribution of death to be COVID-19 related, and the circumstances and likely causes of other deaths.

Finally, COVID-19 has highlighted that care-home residents are relatively invisible in routine data, because we cannot reliably identify care-home residents in routine data.^4^ This has seriously hampered our understanding of the epidemic and its impact, but reflects longer-term neglect of the care sector despite its key role in looking after many of our most vulnerable citizens. In the medium term, we need care-home residence to be accurately recorded to support systematic understanding of the needs of a highly vulnerable population. Similar issues apply to people receiving social care in their own homes, who are also vulnerable and largely invisible in routine data.

## Conclusion

The impact of COVID-19 is concentrated in a small number of care-homes with repeated or sustained outbreaks where the impact has been very large in terms of the numbers of residents, family and staff affected, either directly or by the loss of loved ones. Many care-homes for older people and virtually all other care homes do not yet appear to have had an outbreak. There is therefore considerable risk of further outbreaks with large number of deaths in care-homes if community COVID-19 incidence increases again. Allowing families and friends to visit residents again is important for quality of life, but needs to be balanced against the need to shield residents of care homes in areas where community incidence is high or increasing. Early detection of outbreaks through regular testing, reliable PPE supply, support for infection control, and measures to ensure safe staffing are all likely to be needed to contain the size of established outbreaks.

## Data Availability

Data may be available on request to the corresponding author, subject to NHS Scotland disclosure controls to prevent identification of individuals.

## Acknowledgements

We would like to acknowledge the contribution to data collection and/or comments on the paper from Audrey Pringle, Jenni Strachan, Lindsey Murphy, Louise Wellington, Peter Harrison from the NHS Lothian Health Protection Team; Alison Milne, Dan Clutterbuck and the members of the Enhanced outreach testing team; and Alison McCallum, Director of Public Health.

## Patient and public involvement

There was no patient or public involvement in the analysis.

## Ethics

Analysis was of public health data at care-home level (with no individual or identifiable patient data) in order to understand the evolution of COVID-19 outbreaks in one health board. Separate research ethics review was not required as this work was undertaken under generic approval for the Lothian Research Safe Haven and the NHS Lothian/University of Edinburgh Dataloch partnership agreement. Care Inspectorate data is publicly available at https://www.careinspectorate.com/index.php/statistics-and-analysis/data-and-analysis and is public sector information licensed under the Open Government Licence v3.0.

## Contributorship

The study was conceived by BG, CE, FG, NH, JES, LW and JB. GB, NH, SS, and MT carried out data collection and management, BG, NH, SS, MT, JB and BG carried out data analysis. JB and BG led the drafting of the paper, and all authors contributed to drafting and revision, and approved the submitted paper.

## Competing interests

All authors have completed the ICMJE uniform disclosure form and declare: no support from any organisation for the submitted work; no financial relationships with any organisations that might have an interest in the submitted work in the previous three years, and no other relationships or activities that could appear to have influenced the submitted work.

**Box 1: Initial public health response to COVID-19 in care-homes**

#### February

In the early phases of the COVID epidemic Scotland implemented contact tracing to contain the community spread of the virus. This time was also used to prepare the NHS for an anticipated influx of seriously ill patients requiring hospital care and often intensive care. As well as redesigning patient flows to manage increased infection risk this meant assertively discharging patients who required hospital care with over 900 people discharged from hospital to Scottish care-homes.

#### March

The first positive COVID-19 cases diagnosed in the health board were in early March, mainly in travellers returning from Italy and Spain. As positive cases began to be reported and numbers rapidly increase across Scotland from late March, the Scottish Government and Public Health Scotland (often following Public Health England publications) produced a series of guidance documents. The first document for Nursing Home and Residential Care Home Residents suspending ‘routine visiting’ was issued in mid-March, just as the UK moved from the ‘containment’ to the ‘delay’ phase of the Government’s COVID-19 response. Documents were regularly updated and clarifications made about infection control and late March saw great concern about the availability and distribution of Personal Protective Equipment (PPE) with many reports of scarcity for frontline NHS and particularly social care sector staff. Once contact tracing was abandoned nationally on 13^th^ March, COVID-19 testing resources were directed towards hospitalised patients, and later towards NHS staff. Care home resident testing was available in the board from the second week of March, but testing was initially restricted to only the first few cases to establish the presence of an outbreak with subsequent symptomatic residents assumed to have COVID-19, except in care-homes with large and prolonged outbreaks where wider testing was deployed.

#### April

Similar to other parts of the UK, testing was only extended to care-home staff in early April. As availability of testing improved, Scottish Government specified that all symptomatic care-home residents should be tested from 17th April. Increased responsibility was placed on Directors of Public Health for the management of the epidemic in care-homes, and NHS Boards were required to provide daily updates to Scottish Government. During April, Public Health and Health and Social Care Partnerships increasingly support to care-homes in relation to more systematic testing, infection control, PPE supply and ensuring safe staffing. Availability of resident testing throughout the period was higher than many other areas because the regional virology laboratory had capacity, and a public health outreach time was used to swab residents where care-homes were unable to do so themselves.

**Box 2: Additional measures taken to contain care home outbreaks of COVID-19 in the health board in May and June 2020**

**Extended staff and resident testing**

Nasopharyngeal swabbing and COVID-19 PCR testing for all staff (permanent and agency) and residents in care homes (and other closed settings such as prisons) with new or sustained outbreaks was introduced in late May 2020. A key aim is to identify asymptomatic or low-level symptomatic staff (staff with positive tests often do not have symptoms in the formal case-definition [fever, cough, loss of sense of smell or taste] but about half of ‘asymptomatic’ staff have a range of other low-level symptoms).

**Contact tracing and reinforcing household isolation**

Staff with positive tests are contact traced and advice for self and household isolation reinforced (many live with other care workers).

**Support for the care workers with COVID-19 to self-isolate**

Care workers are a low-paid, marginalised workforce who typically have worse employment conditions than NHS staff in relation to hourly wage and paid sick leave, which makes self and household isolation more challenging. Health Promotion in the health board is working with public health to develop advice and behavioural interventions. In some care homes with ongoing outbreaks in late May, minimal or no sick pay was a clear barrier to staff self-isolating. The health board is supporting rapid access to alternatives to employment-related sick pay via Health and Social Care Partnerships and the Public Health Act (Scotland) 2008, although a barrier is ensuring that this ‘sick pay’ is promptly approved and paid.

#### Support for staffing

Since mid-May 2020, care homes with staffing problems in the face of an outbreak can request support from Health and Social Care Partnerships via access to the Health Board Staff Bank. Bank staff are tested 48 hours before going into any care home.

